# Intensive Care Unit Resource Planning During COVID-19 Emergency at the Regional Level: the Italian case

**DOI:** 10.1101/2020.03.17.20037788

**Authors:** Pietro Hiram Guzzi, Giuseppe Tradigo, Pierangelo Veltri

**Affiliations:** University of Catanzaro

## Abstract

Severe acute respiratory syndrome COVID-19 (SARS-CoV-2) has been declared a worldwide emergency and a pandemic disease by the World Health Organisation (WHO). It started in China in December 2019, and it is currently rapidly spreading throughout Italy, which is now the most affected country after China. There is great attention for the diffusion and evolution of the COVID-19 infection which started from the north (particularly in the Lombardia region) and it is now rapidly affecting other Italian regions. We investigate on the impact of patients hospitalisation in Intensive Care Units (ICUs) at a regional and subregional granularity. We propose a model derived from well-known models in epidemic to estimate the needed number of places in intensive care units. The model will help decision-makers to plan resources in the short and medium-term in order to guarantee appropriate treatments to all patients needing it. We analyse Italian data at regional level up to March 15th aiming to: (i) support health and government decision-makers in gathering rapid and efficient decisions on increasing health structures capacities (in terms of ICU slots) and (ii) define a scalable geographic model to plan emergency and future COVID-19 patients management using reallocating them among health structures. Finally, the here proposed model can be useful in countries where COVID-19 is not yet strongly diffused.

## 1 Introduction

The coronavirus disease COVID-19 was reported in Wuhan (China) on December 2019 [10]. In one month, the surprising diffusion of this virus and a delay on the introduction of severe measures for containing it caused more than 80, 981 confirmed cases and more than 3, 000 deaths [13].

The COVID-19 disease is caused by the SARS-CoV-2 virus that belongs to the Coronaviridæfamily. The Coronaviridæ family contains other viruses that are responsible for respiratory diseases such as the common flu and the complex MERS (middle east respiratory syndrome) and SARS (severe acute respiratory syndrome). Coronavirus family contains many viruses, while only seven are known to be responsible for human diseases (229E, NL63, OC43, HKU1, MERS-CoV, SARS-CoV, and SARS-CoV-2) [9]. One of the main differences between the novel virus and the previous ones is its high spreading rate. Such a rate and the absence of tailored therapies and vaccines determine a relatively high mortality rate that has been controlled by blocking the virus spreading with severe mobility restrictions to the people of the infected regions [13].

At the date of March 15th, while the situation in China seems to be now under control [12], the virus is rapidly growing in other countries, such as Korea, Iran and Italy [2]. Other countries such as the USA, France, Spain and North Europe ones are implementing containment measures. The main problem is related to the exponential diffusion of infections requiring: (i) home quarantine for low symptoms, (ii) hospitalisation for part of them and, (iii) hospitalisation in ICUs requiring respiratory support for severe ones.

From a clinical point of view, patients affected by COVID-19 usually present symptoms similar to common flu, e.g. fever, dry cough, sickness, and breathing problems. A fraction of patients do not need hospitalisation, and the symptoms disappear in a short time (around 1-2 weeks). In some cases, COVID-19 causes severe pneumonia, which requires respiratory support and can lead to death, especially in the presence of co-morbidities such as diabetes or hypertension [3]. Patients with severe pneumonia need to be treated in ICUs with the use of mechanical ventilators [1].

We focus on disease diffusion modelling at a regional scale in Italy and also modelling ICUs Availability throughout Italian health structures. We propose a prediction model at a regional scale that can be adapted and also used at different scales (i.e., countries, regions or even single district) by countries where COVID-19 diffusion is still growing.

## 2 Data analysis and model

We start from the analysis of epidemiological data from Wuhan city (China, Hubey region). As reported in [6, 5] about a third of infected patients became critically ill requiring intensive care unit admission. The lesson learned in China has been used in other countries such as Singapore for preparing a correct strategy for managing people. As noted in [8], preparing ICUs for COVID-19 patients has special requirements such as controlling infection diffusion from patient to medical staff and among patients avoiding the use of ICUs occupied by other patients. Moreover, the evolving scenario related to COVID-19 has required more frequent communication among clinicians.

In Italy^1^ on March 15, we report 24, 747 total cases, 20, 603 people currently affected, 1, 809 deaths and 2, 335 recovered patients. Regarding infected people: 9, 268 are treated in their homes since they do not have severe ill, 9, 663 patients have been hospitalised, and 1, 672 patients have been admitted to ICUs.

To react to the exponential growth of infected patients requiring hospital isation, the Chinese government decided to build a large emergency hospital dedicated to COVID-19 patients in a few days. In Italy, the plan is to improve existing structures by extending the number of ICU resources and beds, as well as using dedicated health structures. E.g., the study reported in [11] focuses on accelerating the process of acquiring and furnish hospitals with assisted breathing devices.Italy has approximately 5, 200 beds in ICUs, which, by law, are designed to be occupied by patients for the 80% at any given time. Also, these are allocated at a regional level proportionally to its population and are usually managed locally (see Table 1).

**Table 1:** Distribution of ICU beds in each region. Such data could increase in the future due to government investments for the emergency.

| **Region** | **Beds** | **Region** | **Beds** |
| --- | --- | --- | --- |
| Piemonte | 320 | Marche | 108 |
| ValleD'Aosta | 15 | Lazio | 590 |
| Lombardia | 1067 | Abruzzo | 73 |
| P.A. Bolzano | 48 | Molise | 30 |
| P.A. Trento | 23 | Campania | 350 |
| Veneto | 498 | Puglia | 210 |
| Friuli Venezia Giulia | 80 | Basilicata | 49 |
| Liguria | 70 | Calabria | 110 |
| Emilia Romagna | 539 | Sicilia | 346 |
| Toscana | 450 | Sardegna | 150 |
| Umbria | 30 | **Italy** | **5156** |

Many of such ICU slots are yet occupied by non-COVID-19 patients while as of March 15 1, 672 beds are occupied by COVID-19 patients. Considering the data progression [4], it is reasonable to predict that the number of ICUs patients will increase. Considering that the average survival time for non-survivor patients is around ten days, there is the need to make decisions about ICU resources. Such decision may regard, for instance, the institution of new ICU beds, the movement of people from one region to another (to improve the overall dislocation of patients by grouping COVID-19 people in a single place). It is trivial that such decision must be based on the correct estimation of ICUs patient, but this estimation is still a matter of discussion [11].

### 2.1 Diffusion Model of SARS-CoV-2

We focus on decision strategy to increase number and structures able to treat COVID-19 patients in intensive units, and thus increasing the number of ICUs. We propose a model able to manage in geographic scale the incoming patients and the ICUs available places. We cover the a need for the development of a predictive model for helping healthcare administrators in managing structure requirements to improve hospitals and patients managements. We extend a compartmental model for epidemiology, and we derive from Italian public data the experimental parameters for simulating the model. Literature contains many mathematical epidemiological models for studying the dynamics of infectious diseases [7]. These models fall in two main classes: deterministic models that are based on differential equations and stochastic models that are based on Markov processes. We here use a discrete-time Markov chain model [14] and we derive the parameters of the model starting from public available data. We use as reference a compartmental model which we adapt from the literature (see Figure 2.1).

**Figure 1:**
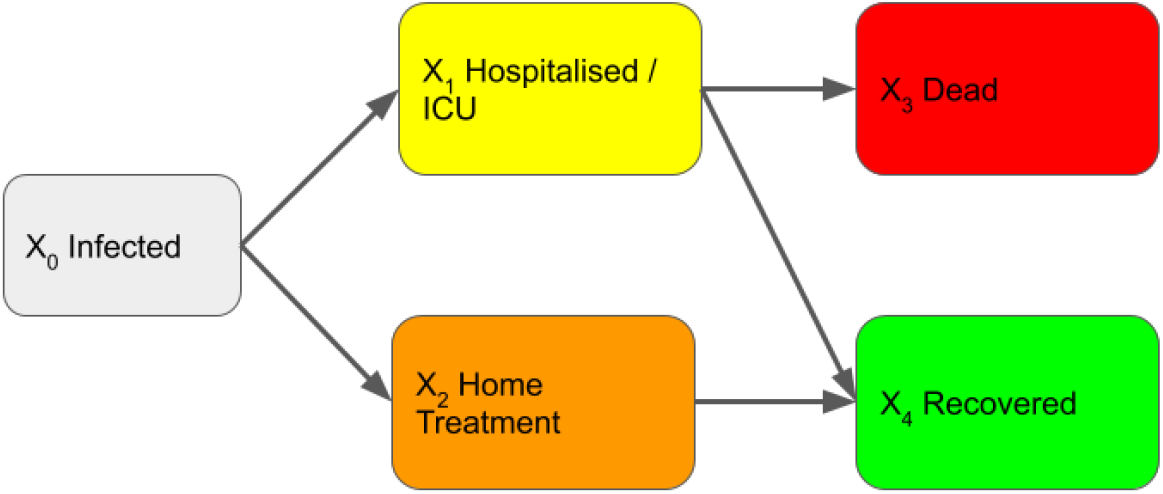
The Compartmental Model. Our model start by considering infected people (*X*_0_). A fraction of infected people presents severe symptoms and they need to be hospitalised and treated in ICUs (*X*_1_). Diversely, some people may be treated at home (*X*_2_) since they do not have severe complications. Treated people has a lethal outcome (*X*_3_) while a hopefully large fraction of people is recovered from disease.

**Figure 2:**
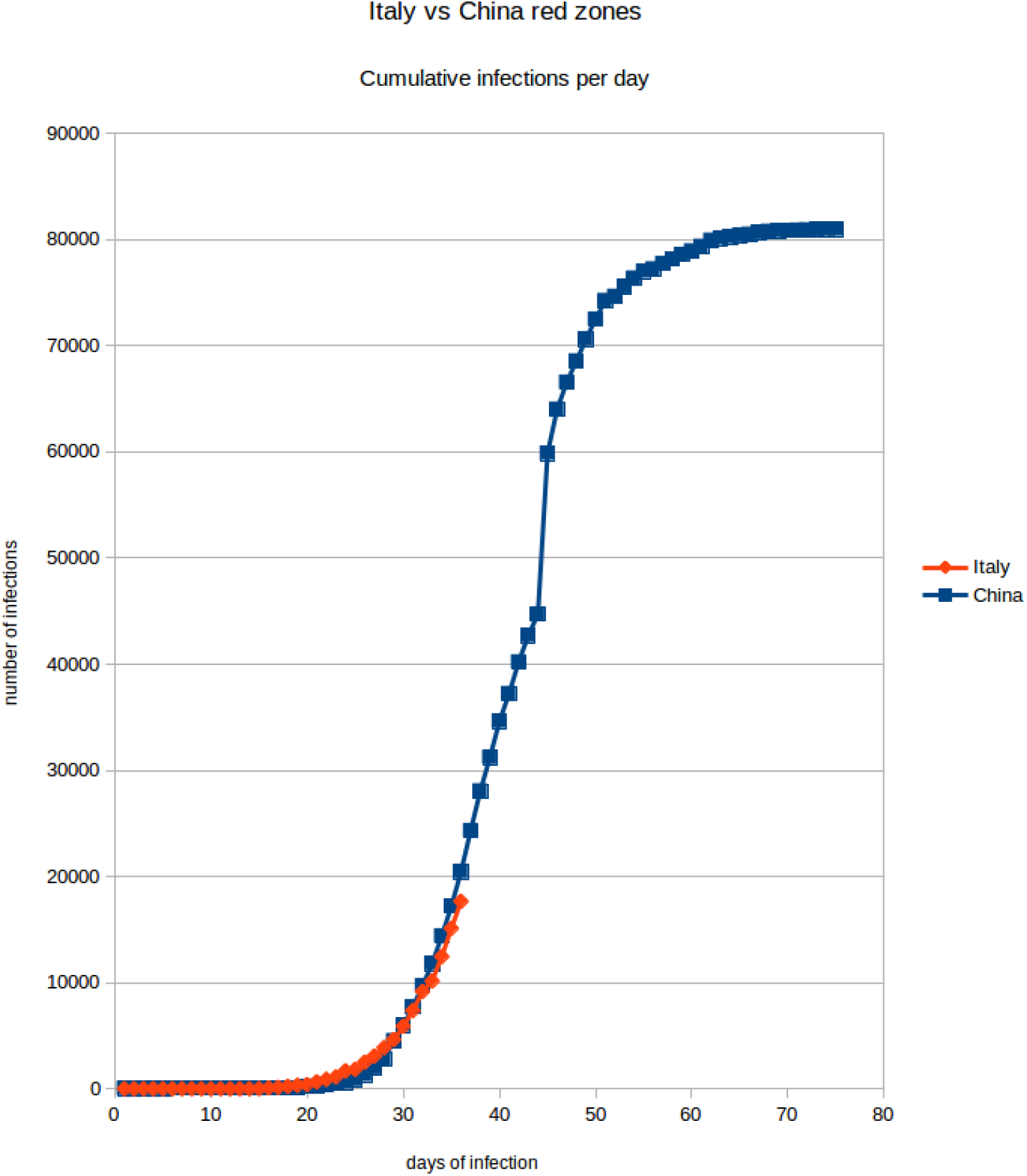
Here the two Italian and Chinese red zones (areas of maximal infection) are compared. On the X axis there are days and on the Y axis there are the total number of cases. The two curves are very similar showing that the initial trend of the infection follows an exponential growth, even though the Chinese government rapidly adopted stringent confinement measures. We can thus expect to observe the same initial infection evolution, before arriving to the logistic portion of the curve.

In figure 2.1 the COVID-19 diffusion is reported both for Italy and China (red zones) at current status. We note that: (i) we can make the hypothesis of similar trend for different countries (including Italy) and that in the logistic phase diseases should be treated even if not in emergency to manage the fraction of patients that require ICUs.

## 3 Regional Scale Geographic distribution

The Italian National Health Service is organised on a national and regional scale. The central government controls the distribution of resources and services are arranged at a regional scale. There are 19 regions and two autonomous provinces (21 total administrative units). Therefore the ICUs is availability is organised at a regional scale, established by each region. Table 1 summarise current ICU beds availability per administrative units. Patients are admitted into the ICUs of its region, without considering other criteria, such as free beds into ICUs of other regions that may be geographically closer. The access is freely guaranteed costs are mapped to citizen with respect to their regions of residence.

This may cause the undesired case that some regions may have many available beds, and other regions do not. This is what is happening in north Italy where restriction plans to reduce citizen mobility has been applied at the regional scale. Figure 3 shows the distribution of total ICU beds versus occupied ICU beds (i.e. in hospitals) for each region in Italy. Figure 3 shows the infected cases for each region.

**Figure 3:**
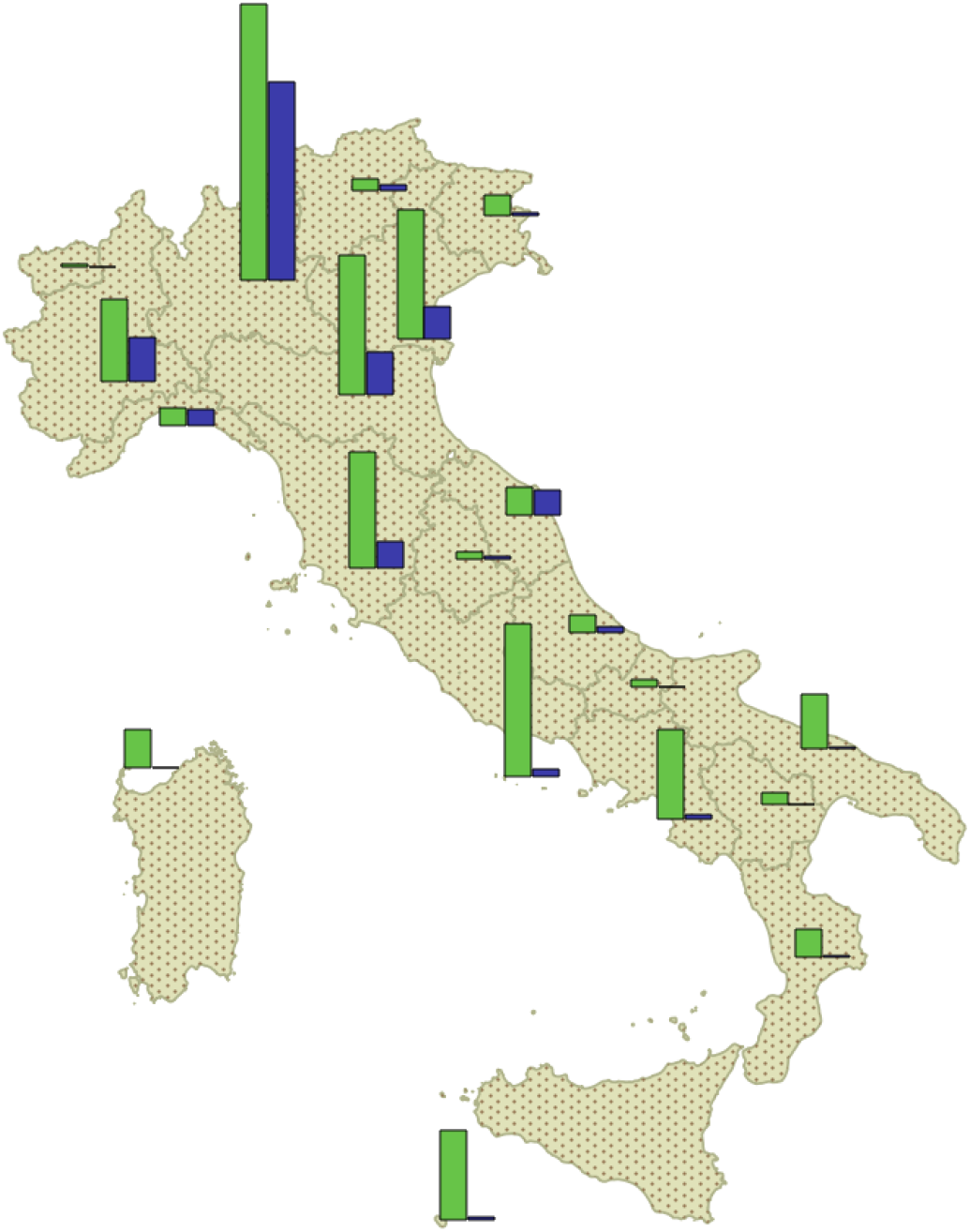
Figure depicts the total number of ICU beds and the number of currently occupied per region. Data is available online on the Italian Civil Protection Department website at https://github.com/pcm-dpc/COVID-19/blob/master/dati-regioni/dpc-covid19-ita-regioni-20200315.csv with licence CC-BY-4.0.

We compare through our model the management of beds in single regions as compartment and the management of places on a nationwide scale (admitting transfers among regions). Our findings suggest that the management of ICUs beds as a whole across regions may improve the overall availability of free beds for COVID-19 patients.

Figure 3 shows the distribution of total ICU beds versus occupied ICU beds (i.e. in hospitals) for each region in Italy. Figure 3 shows the infected cases for each region.

## 4 Model based ICUs Prediction

We want to relate the number of ICU beds with infectious cases. In particular, the target is to predict the number of ICUs bed necessary and required for the amount of infections in a region.

Starting from the representation of infectious diseases trend, we map trend for each Italian region. Then we map and elaborate trend of ICUs bed requirements for infections in each region. This is used to relate infections and ICUs beds (see Figure 4 for Lombardia region). Predicting the number of diseases by using the exponential equation of Figure 4, we may guess the number of infectious in single regions in a short time and thus using such a number to predict the number of ICUs required. These predictions may be used to plan necessity in a short time and also eventually map ICUs non-COVID-19 patients in different regions by reallocating patients in different related structures (hospitals).

**Figure 4:**
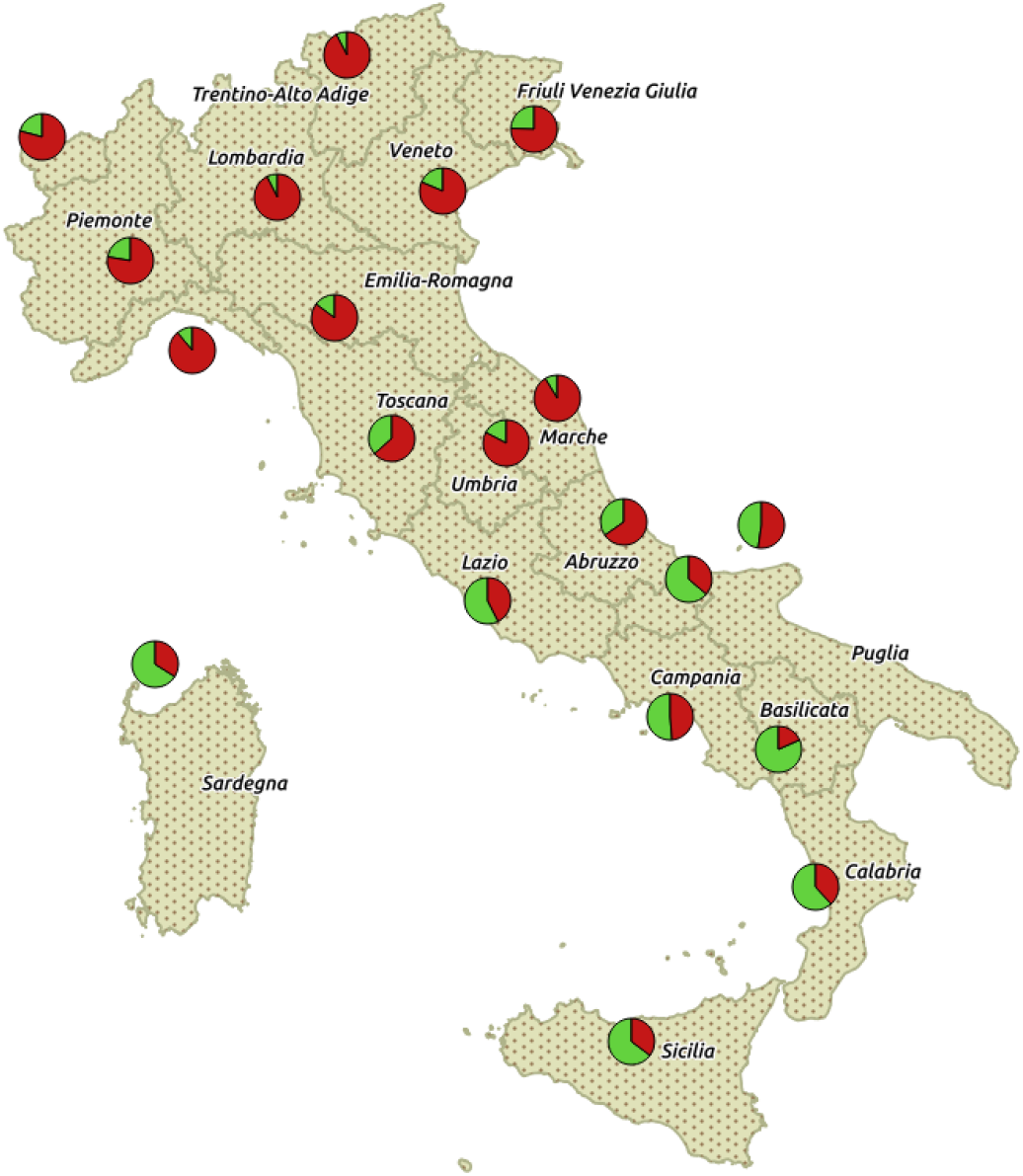
Figure depicts the distribution of COVID-19 infected cases along Italian peninsula. Data is available online on the Italian Civil Protection Department website at https://github.com/pcm-dpc/COVID-19/blob/master/dati-regioni/dpc-covid19-ita-regioni-20200315.csv with licence CC-BY-4.0.

**Figure 5:**
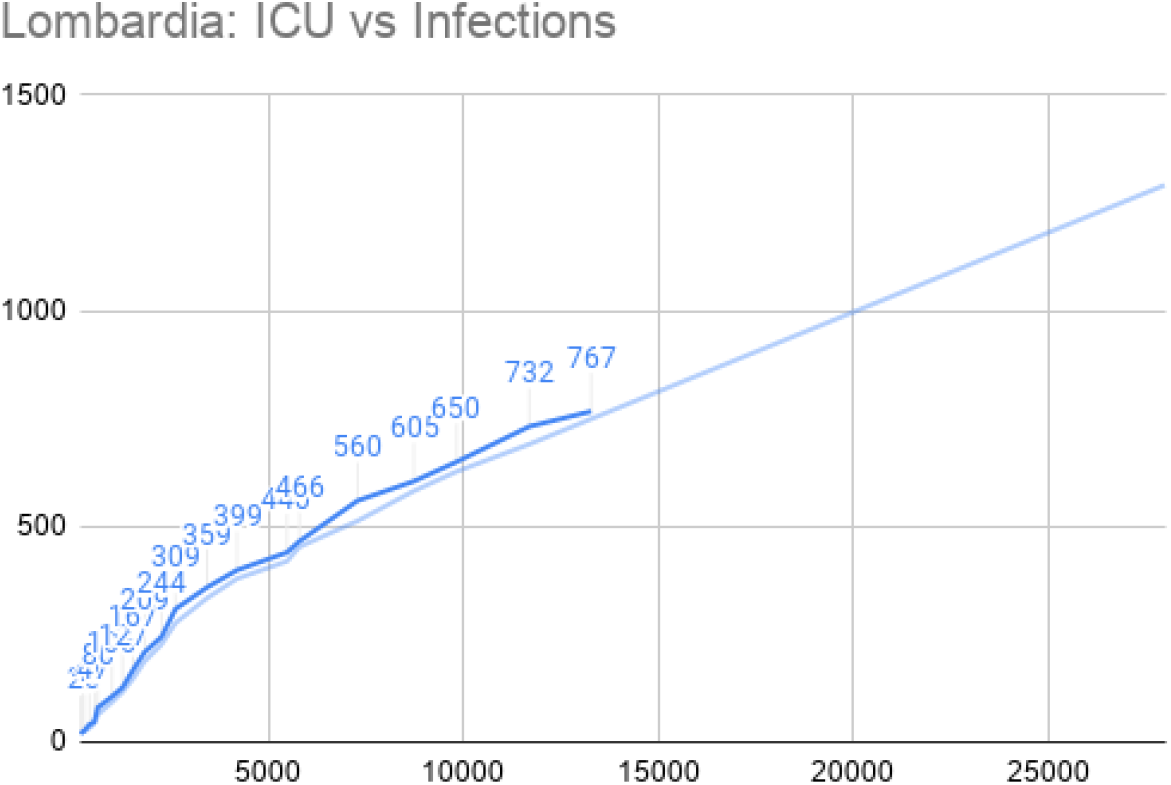
Regional Diffusion of Infections in the Lombardia at the date of March 15th.

**Figure 6:**
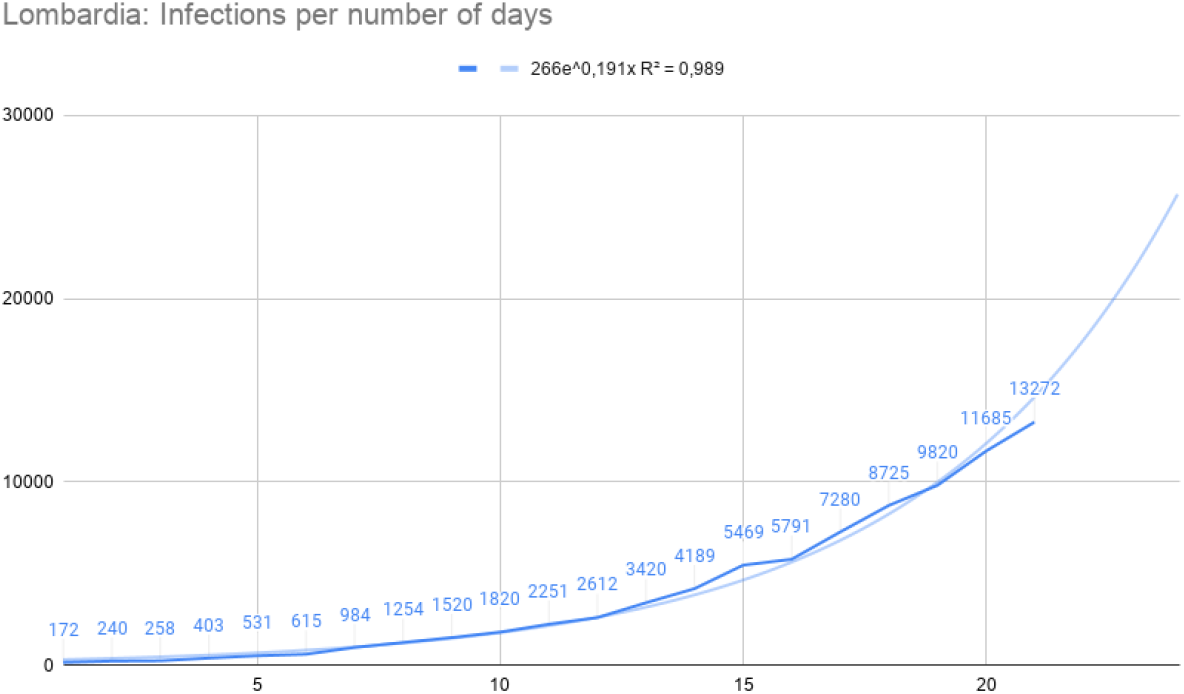
ICU beds prediction graph for the Lombardia region.

**Figure 7:**
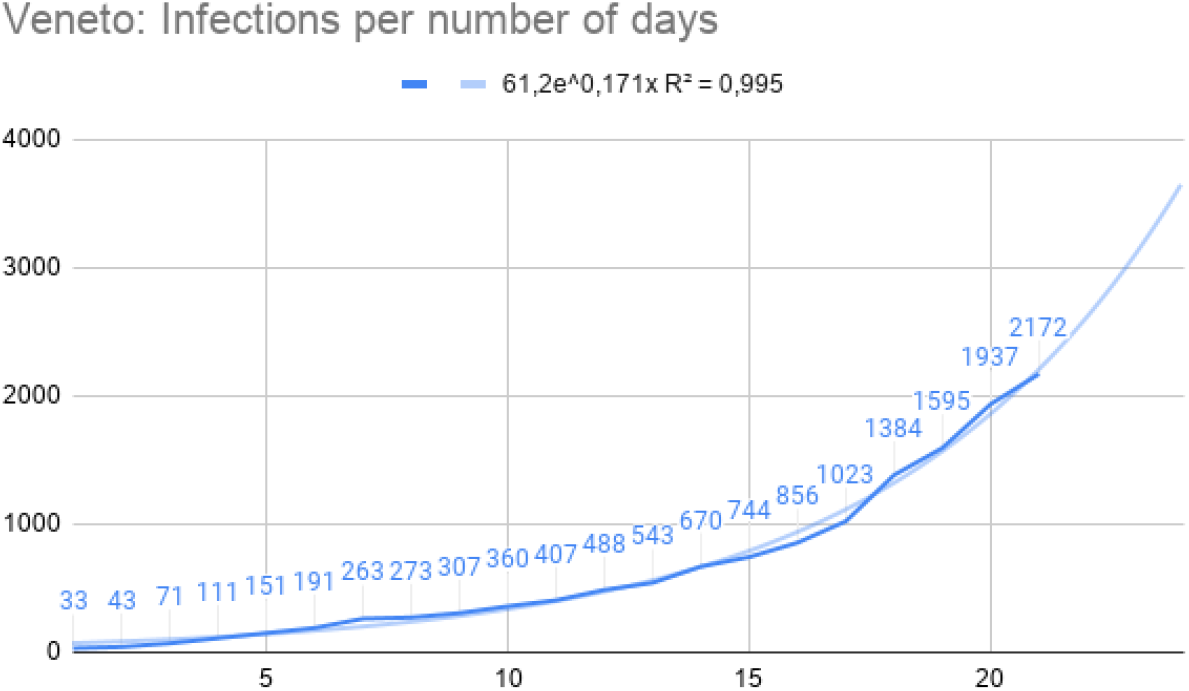
Regional Diffusion of Infections in the Veneto region at the date of March, 15th.

**Figure 8:**
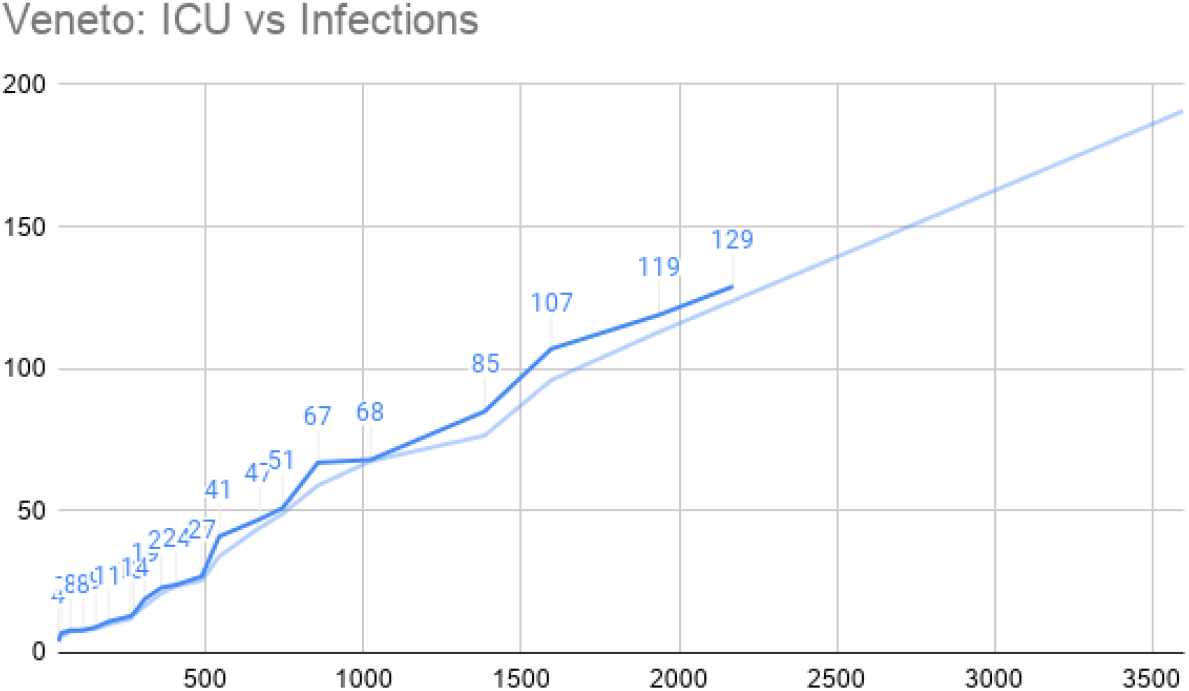
Regional Diffusion of Infections vs ICU beds in Veneto Region at the date of March, 15th.

**Figure 9:**
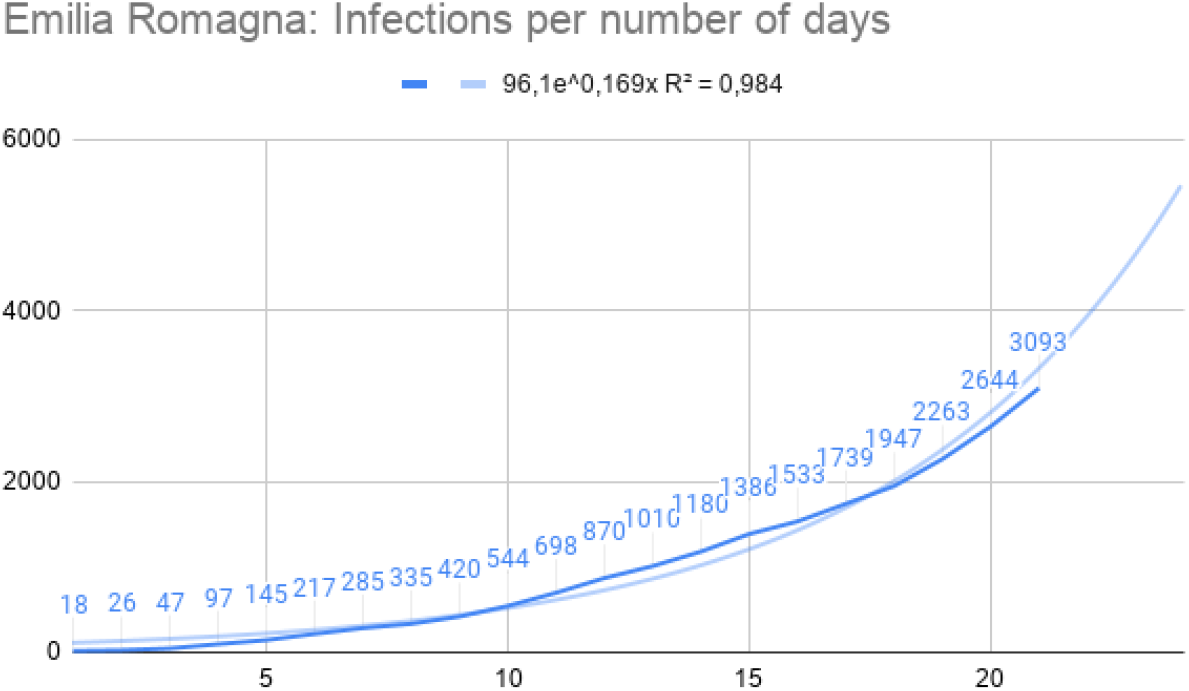
Regional Diffusion of Infections in Emilia Romagna Region at the date of March, 15th.

**Figure 10:**
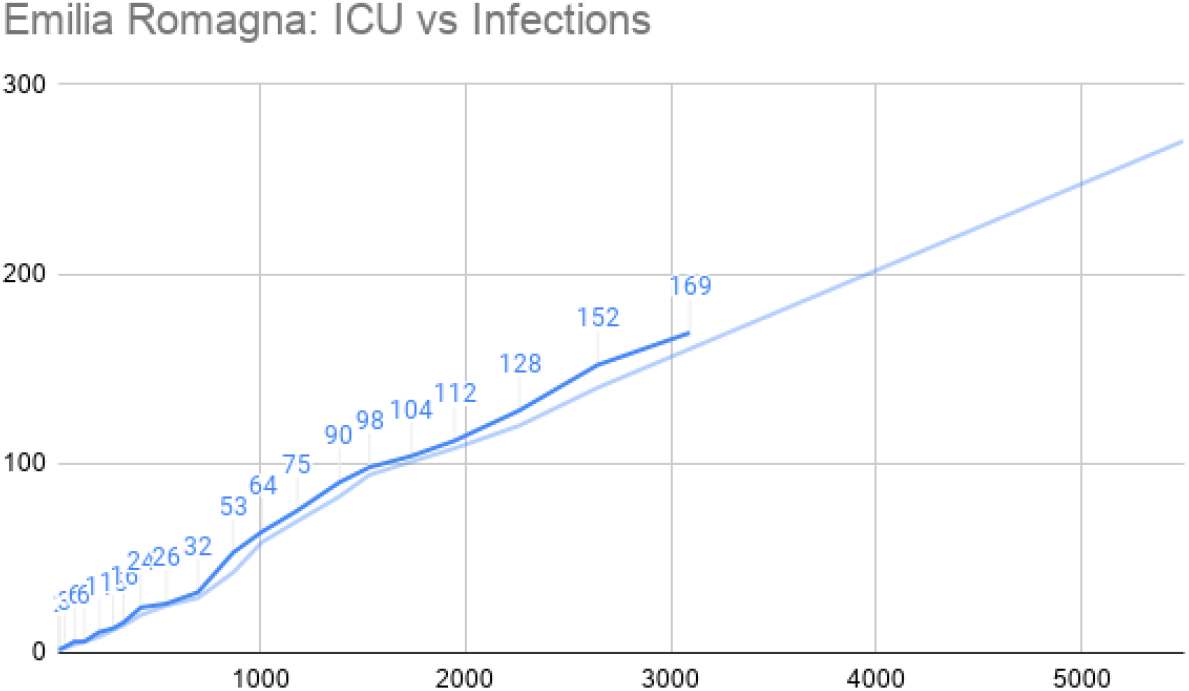
Regional Diffusion of Infections vs ICU beds in Emilia-Romagna Region at the date of March, 15th.

**Figure 11:**
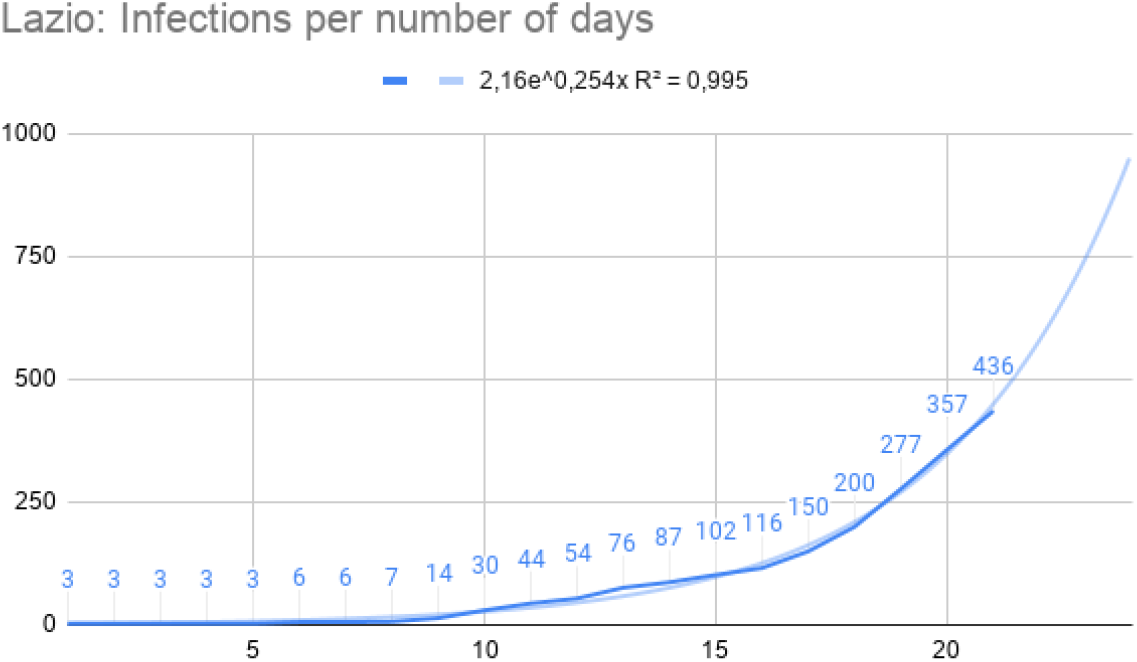
Regional Diffusion of Infections in Lazio Region (Central part of Italy) at the date of March 15th.

**Figure 12:**
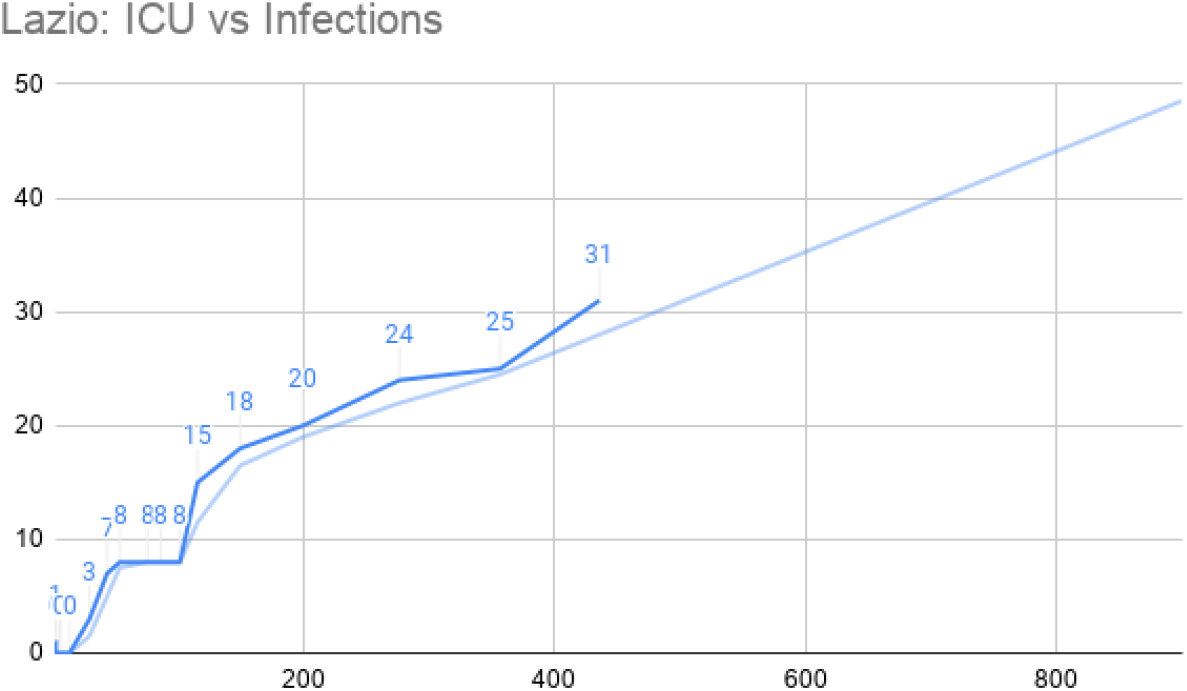
Regional Diffusion of Infections in Lazio Region versus used ICUs at the date of March 15th.

**Figure 13:**
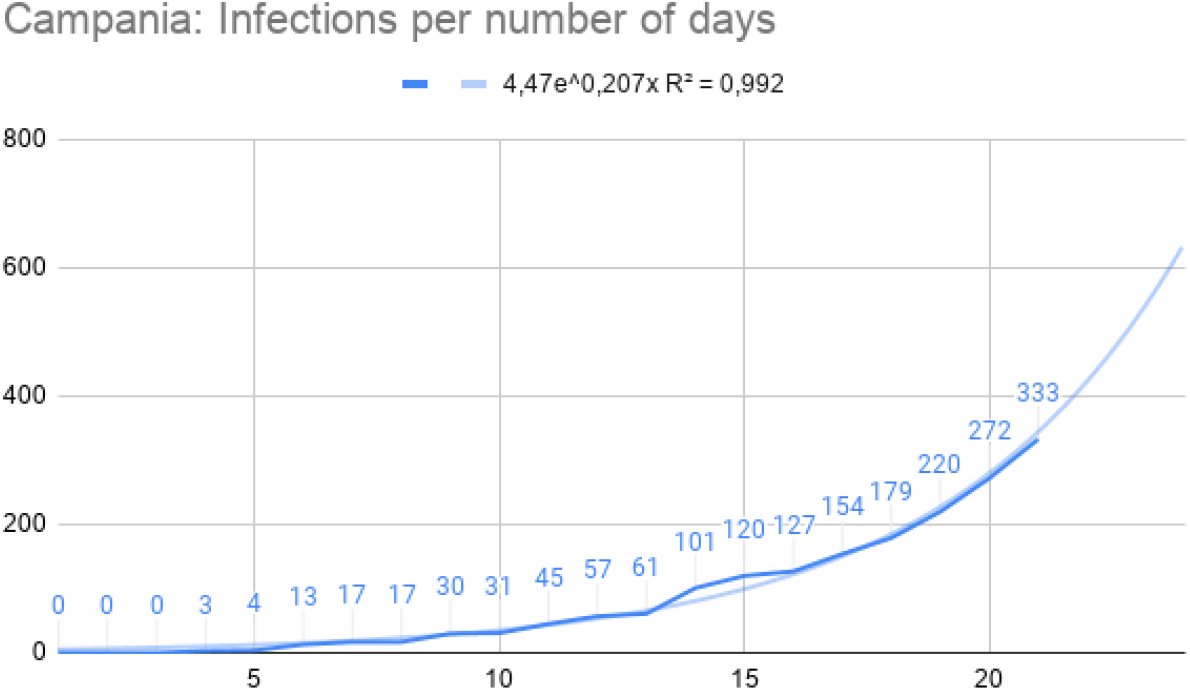
Regional Diffusion of Infections in Campania Region as south italian representative one at the date of March 15th.

**Figure 14:**
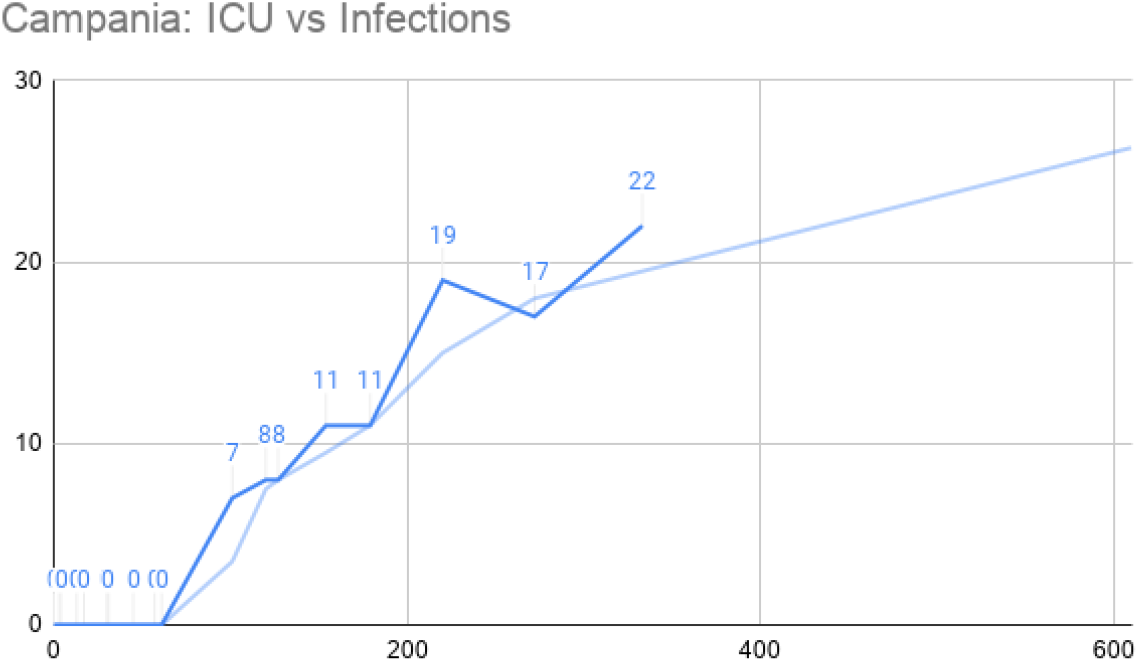
Regional Diffusion of Infections vs number of ICUs used places in Campania Region as south italian representative one at the date of March 15th.

Similarly, for currently most COVID-19 interested Italian regions, trends are reported in Figures 4, 4 for Veneto region, and Figure 4 and 4 for Emilia Romagna region. Lombardia, Veneto and Emilia Romagna are the top three regions with an emergency in terms of ICU beds necessity at the date of March 15th (*saturated regions*). These regions are already starting the re-allocation plans for ICU patients in different regions as well as emergency plans to free ICU beds.

### Predicting ICU for non-saturated regions

Italian regions in the centre and south of Italy, are still living an increasing phase of infection disease with respect to ICU beds capacity (i.e. *non saturated regions*, see Figure 3). We propose to use the predicting model for these regions to early predict saturation phases. In Figures 4 and 4 a diffusion of disease and relative connections with ICUs requirements are reported and refer to a central Italian region, i.e. Lazio, whose capital is Rome. By using both a decisor can calculate the number of infection for some time point in the future and derive the number of predicted ICU beds which will be occupied. Similarly, in Figure 4 we report Campania region situation at March 15th as south of Italy representation. In such case note that the government restrictions rapidly adopted allow a slower diffusion of infectious.

Note that in a similar way we map all data for all the 21 Italian regions.

## 5 Conclusion

The emergency of COVID-19 is related to an aggressive virus that diffuses rapidly and strongly stresses the resistance of health structures. We also think that patients management is strictly related to the availability of hospital structure to manage such kind of diseases that require different and non-standard protocols as use of respiratory devices. We think that by using a scalable predictive model at regional as well as at district granularity may support regional and national government in managing the emergency. We finally claim that such a model can be used in countries where diffusion is still at the beginning as in the US, French, Spain and other European countries as reported in ^2^ where different virus diffusion trajectories are reported for different countries.

## Data Availability

All the data used in this work are publicly available.

## 6 Contributors

GT was responsible for data analysis and statistics, and writing of the manuscript. PHG was responsible for data analysis and writing of the manuscript. PV was responsible for data analysis and writing of the manuscript.

## 8 Acknowledgments

We thank Italian Protezione Civile for freely providing online data thus allowing studies on COVID-19. We thank Prof. Tamer Kahveci from University of Florida for useful suggestions.

## Footnotes

1 http://www.salute.gov.it/portale/nuovocoronavirus/dettaglioContenutiNuovoCoronavirus.jsp

2 https://www.ft.com/content/ff3affea-63c7-11ea-b3f3-fe4680ea68b5

